# A qualitative study of emergency nurses’ perspectives on intranasal ketamine for procedural sedation in children

**DOI:** 10.1101/2024.05.16.24307340

**Authors:** David Wonnacott, Shannon D. Scott, Rachel Flynn, Samina Ali, Everly Van Der Vaart, Naveen Poonai

## Abstract

**Purpose:** There is mounting evidence supporting a role for intranasal (IN) ketamine for procedural sedation in children due to its less invasive delivery. Drug administration and monitoring is largely performed by nurses and clinical uptake requires understanding their perceptions. We explored nursing perspectives of IN ketamine for procedural sedation in children to understand facilitators and barriers and inform institutional guidelines.

**Design and Methods:** From January to February, 2018, we conducted 2 focus groups with 8 registered nurses in a Canadian tertiary care paediatric ED. Following professional transcription, data were analyzed using an inductive qualitative approach.

**Results:** Seven of 8 participants had experience administering IN ketamine to children for procedural sedation. Nurses perceived that IN ketamine had the potential to reduce children’s distress and improve nursing resource use. Perceived barriers included: 1) uncertainty regarding sedation effectiveness and incorporation into institutional sedation protocols, 2) perceptions that IN ketamine produced a relatively lighter, slower-onset, and less titratable sedation, and 3) healthcare providers’ lack of familiarity with IN ketamine and reluctance to change their current approach to sedation.

**Conclusions:** We identified barriers to adoption of IN ketamine such as uncertainty regarding its pharmacodynamic properties, safety, and impact on workflow, along with facilitators such as fewer adverse events and nursing resources, and less procedural distress for children.

**Practice Implications:** Provider education should focus on IN ketamine’s pharmacodynamic properties and development of institutional sedation guidelines that define indications for use, support engagement of child life specialists, and operationalize the type and duration of monitoring requirements.

**CLINICIAN’S CAPSULE:** *What is known about the topic?:*

- Intranasal ketamine is an emerging agent for procedural sedation and analgesia in the emergency department due to ease of administration.

*What did this study ask?:*

- What are paediatric nurses’ perspectives on intranasal ketamine for procedural sedation among children in the emergency department?

*What did this study find?:*

- Nurses identified advantages to intranasal ketamine but expressed considerable uncertainty regarding its pharmacodynamic properties and its incorporation into clinical practice.

*Why does this study matter to clinicians?:*

- Provider education may overcome some uncertainty and should focus on intranasal ketamine’s pharmacodynamic properties and incorporation into institutional sedation protocols.

**Meetings:** Pediatric Academic Societies (Baltimore, Maryland, April 30, 2019); Canadian Association of Emergency Physicians (Halifax, Nova Scotia, May 29, 2019); Canadian Paediatric Society (Toronto, Ontario, June 7, 2019)

## INTRODUCTION

Orthopedic injuries, which often require intravenous (IV) sedation for fracture reduction, comprise more than 10% of emergency department (ED) visits in children (1, 2), making venipuncture extremely common. Local anesthetic can significantly reduce the pain of IV insertion (3), but the procedure is still associated with significant anxiety in children and parents (4). Moreover, IV insertion can be more technically difficult in children because of smaller veins (5) and lack of cooperation, (5, 6), often leading to multiple IV attempts. Among providers, placing IVs in children is associated with lower nursing morale and greater nursing resources (7, 8). Intranasal (IN) medications may obviate the need for distressing IV placement and offer a technically easier and pain-free approach to sedation (9). In the last decade, there has been mounting evidence on the effectiveness of IN ketamine for pain (9, 10), sedation (11), and diagnostic imaging (12). Although IN ketamine has a longer duration of sedation compared to IV ketamine (13), survey data suggest that most parents would accept longer ED stays to avoid IV placement in their children (14). However, nursing perceptions of IN ketamine can greatly inform the logistics of adoption. Our objectives were to identify nursing perceptions of IN ketamine for sedation in children that would otherwise not be apparent from a clinical trial. This information is important to understand the barriers and facilitators to clinical adoption and inform institutional guidelines for its use.

## MATERIALS AND METHODS

### Design

This was a qualitative study using focus groups of paediatric ED nurses. The study adhered to the Consolidated Criteria for Reporting Qualitative Research (COREQ) (15). Approval was obtained from Western University’s Health Sciences Research Ethics Board.

### Setting

The study was conducted at the Children’s Hospital, London Health Sciences Centre. The paediatric ED is staffed by 70, primarily registered nurses, experienced with IV ketamine sedation and IN drug administration, but not specifically IN ketamine, and no institutional guideline for its use exists. IN ketamine was introduced 10 months prior to enrollment as a pilot study of IN ketamine 8 mg/kg for distal forearm reductions in 15 children, age 4-17 years (16).

### Enrollment

Using purposive sampling, participants were recruited using a single email sent to all nurses in our paediatric ED. We included nurses who actively provided clinical care, without exclusions. All eligible respondents were enrolled into one of two, one-hour focus groups, conducted in-person on January 26 and February 2, 2018 in a non-clinical area of the hospital. Participants were given a $50 Visa gift card. Participants were informed that the research was being conducted to explore nursing perceptions and concerns regarding IN ketamine. A male research assistant who was also a family/emergency medicine resident (DW) obtained informed consent, collected data, and conducted the focus groups. The research assistant had no prior relationship with any study participant and this project was an academic requirement. He had no prior experience with IN ketamine and as such, had no prior assumptions or particular interest in the topic beyond the research question. The research assistant’s training involved reviewing literature on conducting focus groups for qualitative studies and several training sessions with a member of the research team experienced in focus groups for qualitative methodology (RF).

### Data Collection

A focus group was chosen to allow participants to discuss open-ended questions, allow for probative questioning, and obtain a broad range of perceptions (17, 18). Data were gathered using a semi-structured interview guide (Appendix) developed by study team members. It was was pilot tested for clarity, validity, comprehension, and length. Interviews explored nursing perceptions around experience, attitudes, perceptions of safety and efficacy of IN ketamine sedation, and willingness to perform procedural sedation without an IV. They were digitally recorded and transcribed verbatim by an accredited court reporter (19). Transcripts were anonymized and analyzed using an inductive qualitative approach described by Nowell et al. (20). The inductive analysis was conducted in three phases: coding, categorizing, and developing themes (21). Coding was conducted independently by two study authors (DW, RF) with disagreements resolved through discussion. Code words reflected the essence of the data leading to ease of recognition as the number of code words increased. Codes were operationally defined so that they could be consistently applied throughout the data and placed into broad categories corresponding to the major unit of analysis. As categories emerged, theoretical properties were defined. Thematic analysis was used to group similar codes into themes.

We also used a deductive approach where we applied Rogers’ Diffusion of Innovation (DOI) theory (22) as a conceptual framework to identify factors that facilitate the adoption of IN ketamine using the following 5 attributes: (i) relative advantage (the degree to which IN ketamine is seen as better than IV ketamine), (ii) compatibility (consistency of IN ketamine with the values, experiences, and needs of health care providers), (iii) complexity (how difficult IN ketamine is to use), (iv) trialability (the extent to which IN ketamine can be tested before a commitment to adopt), and (v) observability (the extent to which IN ketamine provides tangible results) (22). Trustworthiness of the analysis was ensured through a detailed audit trail and peer review of the analytic process. Lincoln and Guba’s 4 criteria guided the trustworthiness of our processes. Throughout the study, an audit trail documented all interpretations as well as an inventory of all data collected. Reflexivity reflection by the research assistant was completed to continually attend to their effect on the collection, analysis and interpretation of the data (23). The analytic process was managed using NVivo 11 (QRS International, Melbourne).

## RESULTS

Participants consisted of 8 female registered nurses accredited by the College of Nurses of Ontario, with a mean (range) of 9 (2-26) years of clinical experience in the paediatric emergency department. There were no dropouts. Seven participants had previous experience administering IN ketamine and monitoring patients. The participant without IN ketamine experience had over 5 years’ experience with other intranasal agents and procedural sedation in children. A summary of themes and subthemes are provided in Table 1.

### Theme: Pharmacodynamic attributes of IN ketamine

This theme focused on nursing perceptions surrounding the clinical attributes of IN ketamine. Participants discussed perceived differences between IN and IV ketamine such as onset, depth, and duration of sedation. Related to DOI theory’s “observability”, participants agreed that IN ketamine produced a relatively lighter, slower-onset, less titratable sedation. For example:

*“I just feel like they don’t really fall asleep [with IN ketamine] the way they do with IV. I just feel like it kind of calms them down and they’re just kind of there on the surface [and] if you do anything, it wakes them up*.”

Participants also expressed that IN ketamine sometimes had a noxious taste, but other adverse events, most prominently emergence phenomenon and vomiting, were less frequent compared to IV ketamine. One nurse reported:

*“They don’t wake up with that terror like they do with the IV ketamine*.*”* Another reported:

*“I think IN ketamine is better because then you are not having to worry about the kid vomiting and aspirating, right? Because they don’t have that side effect post*.*”*

There was a general consensus that IN ketamine’s duration of sedation was longer than IV ketamine. One nurses commented:

*“Just used IN ketamine last week; it took an hour for the patient to return to baseline. She was pretty stoned post*.*”*

Participants identified favorable features of IN administration and expressed that required skills were easily acquired, particularly in comparison to IV placement. For example:

*“And once you’ve seen it done once, then it’s pretty easy to do*.*”*

and

*“I can see a role in something like a rural hospital where they don’t see a lot of paediatric patients and so a lot of times there is a fear of starting an IV because often nurses just aren’t as comfortable, and so I wonder if [with] an IN atomizer, you’d be able to provide that level of comfort, or do simple reductions themselves*.*”*

### Theme: Perceived benefits of IN ketamine

This theme focused on perceived benefits of IN over IV ketamine and proper patient selection. Participants expressed their positive experience with IN ketamine, particularly the opportunity to avoid a painful and distressing experience compared with IV insertion. For example:

*“Some kids are great with getting an IV and then the next one you have to have four staff hold them down. It [IN ketamine] is a better experience for them because the less traumatic they go into sedation, the less traumatic they come out of sedation*.*”*

With respect to parental perceptions, one nurse reported:

*“…not having to put the IV in, because that is almost as traumatic for the parent as it is for the child, and the speed [with] which they recover, and they’re not vomiting, and they’re not green, I think the parents’ perception [of IN ketamine] would better than IV ketamine*.*”*

The importance of patient characteristics on the appropriateness of IN ketamine was a topic of much discussion. Overall, participants felt that IN ketamine was a good anxiolytic and sedative in properly selected patients, with the potential for avoiding distressing IV placement. However, participants identified that patients being too anxious to cooperate obviated many of the advantages of IN ketamine, but that in cooperative patients requiring sedation, it was more useful. Also, most nurses agreed that IV ketamine would be more appropriate for cooperative (older) children, those with unpredictable nasal anatomy, concurrent upper respiratory infection, or who were undergoing relatively more painful or longer procedures. For example:

“*It depends on the volume we’re shoving up their nose too, right? The kids that are heavier, you’re shoving a lot of volume up their nose. They would probably prefer the IV*.*”*

### Theme: Barrier and facilitators to adoption of IN ketamine

This theme focused on nursing attitudes regarding the lack of IV access but also the potential for fewer nursing resources. Related to DOI theory’s “complexity”, participants discussed how IN ketamine may produce discomfort with providing sedation without IV access. Some participants felt that lack of an IV wasn’t a problem. One nurse reported:

*“It’s always airway support anyway, or vomiting, so IV or not, you’re recovering them the exact same way. You’re watching their vitals, you’re watching their oxygen, you’re watching their breathing. You don’t need an IV to do any of that*.*”*

Other nurses expressed being nervous about the lack of an IV, particularly being unable to urgently administer rescue medications. One nurse reported:

*“[Intranasal ketamine sedation] is outside our nursing comfort level because there’s a certain level of control [when] you’ve got them connected to the cardiac monitor and SpO2 and you’ve got a working IV and now all of a sudden, we don’t have IV access and it’s like no control of the unknown*.*”*

Physician preference and resistance to change was identified as a possible barrier to adoption of IN ketamine. Overall, participants felt that IN ketamine had the potential to improve nursing resource utilization by requiring fewer personnel. This reflected the DOI theory’s “relative advantage”. For example:

*“When we use [IN ketamine] for minor procedures like sutures, it also decreases our workload because we’re not having to fight with the child to get them to hold still*.*”*

Another nurse alluded to an advantage of IN ketamine on their workflow:

*“When it decreases parent anxiety, it decreases the child’s anxiety. Their comfort is better, it makes our job easier*.*”*

Some nurses felt that resistance to change and lack of familiarity with IN ketamine among nurses were barriers to adoption. One nurse reported:

*“Us emerg[ency] nurses, we’re not really great for change, and you get used to doing that, and if it works, the theory is: if it’s not broken, why fix it?”*

With respect to physicians:

*“I think the older physicians are very set in their ways, and this is what we do and this is what works, so that’s what we’re going to use*.*”*

### Theme: Impact on ED workflow

This theme focused on a discussion of how IN ketamine sedation would fit into institutional paediatric sedation guidelines. One nurse expressed their uncertainty:

*“Yeah, like the policies around monitoring. I don’t even know if there is one [for IN ketamine]*.*”*

The impact of IN ketamine sedation on the availability of ED beds and monitoring resources was discussed. Regarding DOI theory’s “relative advantage”, participants noted a potential benefit for the ED. For example:

*“[Intranasal ketamine sedation is] usually a shorter procedure. They wake up faster. As soon as it’s a sedation, that nurse is one-on-one. So, the faster the procedure is done,…the better it is for the department*.*”*

### Theme: Uncertainty and novelty

This theme highlighted the lack of personal and institutional familiarity with IN ketamine that permeated the previous themes. At each layer of the discussion, this uncertainty played a significant role in shaping participants’ attitudes. Nurses expressed feelings of uncertainty regarding predictability of IN ketamine’s efficacy, tolerability of IN sprays, and parental perceptions. Related to DOI theory’s “trialability”, there was also uncertainty about how IN ketamine sedation fit within institutional sedation guidelines. Much of this discussion informed DOI theory’s “compatibility”. In particularly, nurses discussed uncertainty around onset and duration of effect and ability to titrate IN ketamine to effect. For example:

*“I find IN [ketamine] definitely less predictable than IV. Everybody comes out of it differently too, but with IN, sometimes it’s longer with some kids depending on how they process it*.*”*

Participants expressed uncertainty about the impact of IN ketamine on patient comfort and compliance, with IN administration believed to be anxiety-sparing for some and anxiety-inducing in others. There was also uncertainty surrounding anticipatory guidance for parents and children, given their lack of personal experience with IN ketamine. For example:

*“I don’t think we’re all there yet, on how to properly educate parents and families on IN ketamine because we don’t really know what’s going to happen*.*”*

Participants also revealed how the novelty of IN ketamine mitigated against traditional teaching that conveyed knowledge from more to less experienced nurses. One nurse recounted:

*“If I have never done something before and it’s new, I’m always concerned. And I always go to a senior nurse to say, “have you done this before and what do you think about it?” and there was no one to do that with because no one had done that before*.*”*

## DISCUSSION

IN ketamine is an emerging agent for sedation and analgesia in the ED (24). This qualitative study of paediatric ED nurses highlighted their perceptions on IN ketamine regarding impact on patients, providers, and resources. It also highlighted important areas of uncertainty as potential barriers to adoption of IN ketamine. Our findings suggest that diffusion of this innovation into practice may benefit from provider education on IN ketamine’s pharmacodynamic properties, proper patient selection, and strategies to optimize compliance, in addition to developing institutional guidelines.

Nurses expressed uncertainty regarding incorporating IN ketamine into institutional sedation guidelines. The protocolization of paediatric pain management and sedation in hospitals has been associated with “improvements in team function and patient care” among interprofessional critical care teams, including nurses (25). This suggests that IN ketamine may be more easily adopted if institutional guidelines were developed for administration, dosing, and monitoring requirements.

Uncertainty regarding the pharmacodynamic characteristics of IN ketamine is not unexpected given the limited number of paediatric trials and clinical experience. In contrast to IV ketamine’s predictable onset and duration of action, roughly one minute and 5-10 minutes, respectively (26), IN ketamine’s onset and duration of sedation ranges widely from 4-12 and 7-69 minutes, respectively (13), making optimal timing of the procedure, monitoring, and discharge difficult to predict. Nursing experience was limited to a pilot study with inconsistent findings of parameters such as efficacy and adverse effects. This was reflected in conflicting perceptions among nurses regarding IN ketamine’s duration sedative action, with at one nurse reporting that IN ketamine administration is usually a “shorter procedure” while another reported that in some children, recovery “is longer…depending on how they process it”.

DOI theory (22) identified attributes of IN ketamine promoting successful adoption and highlighted aspects that may mitigate its use. With respect to “observability” (22), nurses highlighted clear benefits such as avoiding the need to restrain a child for IV placement but also potential disadvantages such as a lighter level of sedation. In the trial which provided local experience with IN ketamine, only 4/7 (57%) of children were adequately sedated. Similarly, a systematic review of 23 trials found that only 200/371 (54%) of children were adequately sedated for IV insertion (27), an often distressing procedure in children (6, 28). A lighter level of sedation is problematic because additional doses of IN ketamine would result in increased time to reach clinical effect.

Regarding “relative advantage” (22), nurses identified favorable features of IN ketamine, including a perception of fewer adverse events, less nursing resources required for placement of an IV, and the opportunity to avoid a distressing procedure for patients and families. The most frequent adverse events with IN ketamine are vomiting and emergence agitation, seen in 8-10% and 6-11% of children, respectively (13, 27), comparable to IV ketamine, (8.4% and 7.6%, respectively) (29). With respect to serious adverse events, 2 systematic reviews of IN ketamine (n=1944) reported only 2 cases (0.1%) of airway and respiratory events, compared to 3.9% with IV ketamine (30). In contrast, the pilot study found adverse events in 4/7 (57%) of children, likely due to higher doses (16). Not all participants may have been privy to all adverse events, some of which (eg. nausea and dizziness), may not have been readily apparent.

Pertaining to “compatibility” (22), nurses expressed uncertainty regarding their ability to provide accurate anticipatory guidance to parents, being able to ask senior colleagues for help, and delivering emergent rescue therapies without IV access. Through discussion, it became clear that paediatric emergency nurses viewed these and other tasks as central to their role in the ED and suggests that widespread adoption may depend on comprehensive education strategies.

Regarding “trialability” (22), nurses reported that resistance to change on the part of providers and uncertainty regarding how to incorporate IN ketamine into institutional sedation guidelines were potential barriers to implementation. Interestingly, implementation of a paediatric pain and sedation protocol in a paediatric cardiac intensive care unit was associated with a greater degree of comfort and health care team functioning (25). In our ED, nurses did not receive formal training on IN drug administration and were guided only by a research protocol for a pilot study of IN ketamine (16). This underscores the importance of developing institutional sedation guidelines for appropriate and safe administration of IN.

In terms of “complexity” (22), nurses highlighted the importance of proper patient selection. IN administration of narcotics by registered nurses is permitted under the College of Nurses of Ontario and the technique requires brief training. In our ED, all nurses had experience administering IN drugs such as midazolam and fentanyl. IN ketamine is administered similarly but requires a greater number of sprays. Perhaps for this reason, nurses perceived that patient cooperation was important and that for older, more cooperative children undergoing longer procedures, IV ketamine would be more appropriate. IN ketamine appears to be most effective for procedural sedation at doses ≥ 7 mg/kg (13, 16). Using the supplied Canadian formulary version of 50 mg/mL and an optimal IN volume of 0.5 mL per nare (31), older (and heavier) patients would require many pairs of sprays. In contrast, younger children, in whom IV insertion can be technically more challenging (5, 6), require a fewer number of sprays. Many Canadian paediatric EDs are staffed by child life specialists, who provide anticipatory guidance and can support children through IN administration (32). When our focus groups were conducted, staffing of child life specialists was limited and nurses did not witness their ability to facilitate IN ketamine administration. In addition to child life specialists, quality improvement initiatives such as The Comfort Promise outline approaches on proper position of young children to reduce procedural distress (33). Although high dose IN ketamine is reportedly well tolerated by children (12), the aforementioned strategies should feature prominently within institutional sedation guidelines.

There was no consensus among nurses regarding their comfort in providing sedation without IV access. Given that adverse events which would require IV access (hypersensitivity reactions and respiratory compromise) affect less than 1% of children sedated with IV ketamine (34) and did not occur in their experience, nurses may not have experienced an adverse event requiring IV access. Nevertheless, adoption of IN ketamine will require education surrounding adverse events and selecting low risk patients. Future studies should specifically explore this concern as it may be an important limitation in achieving provider support for IN ketamine.

## LIMITATIONS

The interviewer was mindful that his role as a physician influenced both his perceptions of IN ketamine, his perceptions of nurses’ responsibilities at the bedside, and the priorities of sedation in the ED, however, being a physician trainee may have shaped the collection and analysis of the data. We did not explore nursing perceptions regarding IN ketamine for analgesia despite mounting evidence that IN ketamine is an effective analgesic in children (35, 36). Given its reduced sedative efficacy compared to IV ketamine, it may be more likely that IN ketamine will find a niche as a non-opioid analgesic. Finally, email recruitment may have selected for nurses who were more willing to share stronger opinions given their experience with IN ketamine. Although this approach yielded a firsthand understanding of nursing perceptions, it may have excluded nurses with more paediatric sedation experience, albeit not with IN ketamine, thereby narrowing the context of the discussion. A wider recruitment strategy enrolling more nurses may have led to a richer discussion that was contextualized in greater experience, thereby highlighting additional facilitators as well as barriers to adoption.

## CONCLUSIONS

Nurses harbor considerable uncertainty regarding IN ketamine. Educational strategies may help overcome this and should focus on IN ketamine’s pharmacodynamic properties to support proper patient selection and indications and provision of anticipatory guidance to patients. Institutional sedation guidelines should define indications for use, support involvement of child life specialists, and specify ED monitoring requirements.

## Data Availability

All data produced in the present work are contained in the manuscript

## REFERENCES

1. Chamberlain JM, Patel KM, Pollack MM, Brayer A, Macias CG, Okada P, et al. Recalibration of the pediatric risk of admission score using a multi-institutional sample. Ann Emerg Med. 2004;43(4):461–8.

2. Hayden JC, Breatnach C, Doherty DR, Healy M, Howlett MM, Gallagher PJ, et al. Efficacy of α2-agonists for sedation in pediatric critical care: a systematic review. Pediatr Crit Care Med. 2016;17(2):e66–75.

3. Cozzi G, Valerio P, Kenedy R. A narrative review with practical advice on how to decrease pain and distress during venepuncture and peripheral intravenous cannulation. Acta Paediatr Scand. 2021;110(2):423–32.

4. Kassi S, Ali S, Scott SD, Hartling L. Procedural pain in children: a qualitative study of caregiver experiences and information needs. BMC Pediatr. 2018;18(1):324.

5. Dychter SS, Gold DA, Carson D, Haller M. Intravenous therapy: a review of complications and economic considerations of peripheral access. J Infus Nurs. 2012;35(2):84–91.

6. Cummings EA, Reid GJ, Finley GA, McGrath PJ, Ritchie JA. Prevalence and source of pain in pediatric inpatients. Pain. 1996;68(1):25–31.

7. Papa A, Morgan R, Zempsky WT. Competency, compassion and contentment: nurses’ attitudes toward pain associated with peripheral venous access in pediatric patients. American Pain Society Annual Meeting. 2008;Abstract 8.

8. Nagy S. A comparison of the effects of patients’ pain on nurses working in burns and neonatal intensive care units. J Adv Nurs. 1998;27(2):335–40.

9. Graudins A, Meek R, Egerton-Warburton D, Oakley E, Seith R. The PICHFORK (Pain in Children Fentanyl or Ketamine) trial: a randomized controlled trial comparing intranasal ketamine and fentanyl for the relief of moderate to severe pain in children with limb injuries. Ann Emerg Med. 2015;65(3):248–54.e1.

10. Nielsen BN, Friis SM, Romsing J, et al. Intranasal sufentanil/ketamine analgesia in children. Paediatr Anaesth. 2014;24(2):170–80.

11. Bilgen S, Köner Ö, Karacay S, Sancar NK, Kaspar EC, Sözübir S. Effect of ketamine versus alfentanil following midazolam in preventing emergence agitation in children after sevoflurane anaesthesia: A prospective randomized clinical trial. J Int Med Res. 2014;42(6):1262–71.

12. Ibrahim M. A prospective, randomized, double blinded comparison of intranasal dexmedetomodine vs intranasal ketamine in combination with intravenous midazolam for procedural sedation in school aged children undergoing MRI. Anesth Essays Res. 2014;8(2):179–86.

13. Poonai N, Canton K, Ali S, et al. Intranasal ketamine for procedural sedation and analgesia in children: a systematic review. PLOS ONE. 2017;12(3).

14. Walsh BM, Bartfield JM. Survey of parental willingness to pay and willingness to stay for “painless” intravenous catheter placement. Pediatr Emerg Care. 2006;22(11):699–703.

15. Tong A, Saintsbury P, Craig J. Consolidated criteria for reporting qualitative research (COREQ): a 32-item checklist for interviews and focus groups. International journal for quality in health care: journal of the International Society for Quality in Health Care / ISQua. 2007;19(6):349–57.

16. Poonai N, Canton K, Elsie S, Joubert G, Shah A, Rieder M. Intranasal ketamine for procedural sedation in children: a randomized controlled pilot study. Ann Emerg Med. 2017;70:S109.

17. Duggleby W. What about focus group interaction data? Qual Health Res. 2005;15:832–40.

18. Kitzinger J. The methodology of focus groups: The importance of interaction between research participants. Soc Health Illness. 1994;16:103–19.

19. Scott SD, Sharpe H, O’Leary K, Dehaeke U, Hindmarsh K, Moore JG, et al. Court reporters: A viable solution for the challenges of focus group data collection?. Qual Health Res. 2009;19(1):140–6.

20. Nowell LS, Norris JM, White DE, Moules NJ. Thematic analysis: Striving to meet the trustworthiness criteria. Int J Qual Methods. 2017;16(1):1–13.

21. Morse M, Field A. Qualitative Research Methods for Health Professionals. Second ed: Sage Publications, Thousand Oaks 1995.

22. Sanson-Fisher RW. Diffusion of innovation theory for clinical change. Med J Aust. 2004;180:S55–6.

23. Lincoln Y, Guba E. Naturalistic inquiry. Newbury Park CA: Sage; 1985.

24. Poonai N, Spohn J, Vandermeer B, et al. Intranasal dexmedetomidine for anxiety-provoking procedures in children: a systematic review and meta-analysis. Pediatrics. 2020;145(1):e20191623.

25. Staveski SL, Wu M, Tesoro TM, Roth SJ, Cisco MJ. Interprofessional team’s perception of care delivery after implementation of a pediatric pain and sedation protocol Crit Care Nurse. 2017;37(3):66– 76.

26. Green SM, Roback MG, Kennedy RM, Krauss B. Clinical Practice Guideline for Emergency Department Ketamine Dissociative Sedation: 2011 Update. Ann Emerg Med. 2011;57(5):449–61.

27. Poonai N, Canton K, Ali S, Hendrikx S, Shah A, Miller M, et al. Intranasal ketamine for anesthetic premedication in children: a systematic review. Pain Management. 2018;8(6):495–503.

28. Ortiz MI, López-Zarco M, Arreola-Bautista EJ. Procedural pain and anxiety in paediatric patients in a Mexican emergency department. J Adv Nurs. 2012;68(12):2700–9.

29. Green SM, Emergency Dept Ketamine M-A, Emergency Department Ketamine Meta-Analysis Study G. Predictors of emesis and recovery agitation with emergency department ketamine sedation: an individual-patient data meta-analysis of 8,282 children. Ann Emerg Med. 2009;54(2):171–80.e4.

30. Green SM, Emergency Dept Ketamine M-A, Emergency Department Ketamine Meta-Analysis Study G. Predictors of airway and respiratory adverse events with ketamine sedation in the emergency department: an individual-patient data meta-analysis of 8,282 children. Ann Emerg Med. 2009;54(2):158–68.e4.

31. Poonai N, Coriolano K, Klassen TP, et al., On behalf of KidsCAN PERC iPCT-SPOR (Innovative Paediatric Clinical Trials – Strategy for Patient Oriented Research) Ketodex Study Team. Adaptive randomised controlled non-inferiority multicentre trial (the Ketodex Trial) on intranasal dexmedetomidine plus ketamine for procedural sedation in children: study protocol. BMJ Open. 2020;10:e041319.

32. American Academy of Pediatrics, Committee on Hospital Care. Child Life Services. Pediatrics. 2020;106(5):1156–9.

33. Friedrichsdorf SJ, Eull D, Weidner C, Postier A. A hospital-wide initiative to eliminate or reduce needle pain in children using lean methodology. Pain Reports. 2018;3:1–11.

34. Bhatt M, Johnson DW, Chan J, et al., On behalf of the Sedation Safety Study Group of Pediatric Emergency Research Canada (PERC). Risk factors for adverse events in emergency department procedural sedation for children. JAMA Pediatr. 2017;171(10):957–64.

35. Frey TM, Florin TA, Caruso M, Zhang N, Zhang Y, Mittiga MR. Effect of intranasal ketamine vs fentanyl on pain reduction for extremity injuries in children: the PRIME randomized clinical trial. JAMA Pediatr. 2019;173(2):140–6.

36. Reynolds SL, Byrant KK, Studnek JR, et al. Randomized controlled feasibility trial of intranasal ketamine compared to intranasal fentanyl for analgesia in children with suspected extremity fractures. Acad Emerg Med. 2017;24(12):1430–40.

